# Inflammatory bowel disease and risk of more than 1500 comorbidities: A disease-wide pre- and post-diagnostic phenomic association study

**DOI:** 10.1101/2024.02.14.24302206

**Authors:** Anthony C Ebert, Rahma Elmahdi, Bram Verstockt, Martin Bøgsted, Gry Poulsen, Aleksejs Sazonovs, Charlie W Lees, Tine Jess

## Abstract

**Introduction:** Inflammatory bowel disease (IBD) is associated with various extra-intestinal manifestations. We aim to identify comorbidities in IBD and the timing of their development to provide valuable insight into the mechanisms underlying IBD.

**Methods:** We conducted a population– and disease-wide phenomic association study in IBD, using *>*6 million ICD-10 coded healthcare contacts from 10 years before and up-to 17 years after IBD diagnosis to investigate associations with 1583 comorbidities. To explore diseases with potential aetiological significance, we compared association in the pre-diagnostic and the post-diagnostic periods. We corrected also for multiple-testing. These estimates were validated with publically available data from Finland.

**Results:** We identified 312 significant associations with 125 appearing before diagnosis. Risk of immune-mediated diseases and extra-intestinal manifestations was increased up to 10 years prior to IBD diagnosis, such as psoriasis (OR_CD_: 2.58 95% CI: [2.00-3.31]; OR_UC_: 1.54 [1.26-1.88]) and reactive arthropathies (OR_CD_: 2.07 [1.42-2.96]; OR_UC_: 1.42 [1.08-1.84]). Risk of cardiometabolic and neuropsychological disorders was increased both pre– and post-diagnostically. Potential treatment sequelae, such as osteoporosis (HR_CD_: 2.56 [2.30-2.86]; HR_UC_: 1.92 [1.79-2.07]) were primarily seen post-diagnostically. In only 15.7% (44/281) and 11.4% (35/301) of comorbidities in CD and UC respectively did we observe heterogeneity between Denmark and Finland.

**Conclusion:** Findings demonstrate that IBD is a multisystemic disease, particularly manifesting with metabolic, immune, and neuropsychological disorders, up-to 10 years prior to diagnosis. We find evidence for the generality of these findings with an international comparison. Diseases of etiological interest warrant further investigation.

**STUDY HIGHLIGHTS:** *WHAT IS KNOWN:*

- IBD is strongly associated with other diseases
- IBD has a complex etiology

*WHAT IS NEW HERE:*

- IBD appears to be a multiorgan systemic disease not confined to the gut
- Associations between IBD and non-digestive disorders are present up to 10 years prior to diagnosis either reflecting a prolonged pre-diagnostic phase or common etiologies
- These findings are validated in an international comparison

## 1 INTRODUCTION

The inflammatory bowel diseases (IBD), Crohn’s disease (CD) and ulcerative colitis (UC), are chronic intestinal conditions with a multifactorial etiology. Identified genetic risk loci ^1;2^ may —in combination with environmental factors, such as diet, smoking, exercise, and exposure to infectious agents—be implicated in disease development ^3^ through multiple mechanisms, including epigenetic modifications, alterations in the gut microbiota, and immune dysregulation ^4^. To complicate the aetiological understanding of IBD further, between 6-50% of patients develop extra-intestinal manifestations (EIMs), including erythema nodosum, uveitis, primary sclerosing cholangitis, and autoimmune hepatitis ^5^, or immune-mediated inflammatory diseases (IMIDs), including psoriasis and ankylosing spondylitis, which may share common inflammatory pathways, genetic associations ^6^, and response to anti-inflammatory therapies including those targeting TNF, IL12/23, and JAK inhibition. Emerging evidence also suggests that neuropsychiatric diseases ^7^ and cardiovascular diseases occur with increased frequency in IBD ^8^, and additional diseases may occur either due to chronic inflammation, altered immunity and/or treatment with corticosteroids and immunosuppression ^9^.

Hence, our understanding of IBD as a single-organ disease is challenged, while evidence of the involvement of other organs is still fragmented. This creates limitations for our aetiological understanding of IBD and the ability to optimize disease phenotyping and tailor treatment. ^10^ In other IMIDs, such as rheumatoid arthritis and psoriasis, there in increasing systematic investigation into associated comorbidities, primarily those manifesting following IMID diagnosis. This has not only enhanced the understanding of disease etiology of diseases such as rheumatoid arthritis and psoriasis but has also revealed that co-morbidities lead to increased cost of care, poorer medical outcomes, ^11;12^ and reduced emotional functioning ^13^. With a compounding prevalence of IBD worldwide and an increasing average age of IBD patients ^14^, the co-morbid burden is only expected to increase further, and a greater understanding of the interrelationship of IBD and comorbid diseases is urgently needed.

Using Danish nationwide healthcare data on all disease categories, as defined by the World Health Organization’s 10th revision of the International Classification of Diseases (ICD-10), we aimed to undertake the first population– and disease-wide association study in IBD exploring all pre-diagnostic comorbidities from 10 to 1 year prior to IBD, as well as all post-diagnostic comorbidities observed from date of IBD diagnosis and up to 17 years after.

## 2 METHODS

### 2.1 Data and study population

The Danish National Patient Register (LPR) covers the entire population of Denmark with comprehensive data from 1977^15^. These data record hospital contacts with associated diagnostic codes including outpatient contacts from 1995 and contacts from private hospitals from 2002. IBD and all other diseases are defined with reference to the World Health Organization’s 10th revision of the International Classification of Diseases (ICD-10) ^16^. The LPR data are linked to the Danish Civil Registration System (CPR) ^17^ by a unique personal identification number to verify and obtain data on age, sex, vital statistics, and residence.

The study cohort comprises all Danish residents with a new IBD diagnosis in the period from 1 January 2005 to 31 December 2021. IBD cases are identified with reference to the LPR with the following criteria, as established in a previous study ^18^, at least two in-or outpatient contacts within a two-year period, with associated (primary or secondary) IBD diagnostic codes (CD: ICD-8 563.01, 563.02, 563.08, 563.09 and ICD-10: K50; UC: ICD-8 563.19, 569.04 and ICD-10 codes K51). Patients with an earlier in-or out-patient contact with an IBD diagnostic code were excluded. A case is defined as either UC or CD based on the most recent diagnostic code and the date of first recorded diagnosis is termed the index-date. To allow adequate time to identify pre-diagnostic comorbidities and to improve identification of only newly diagnosed IBD cases, the IBD patients included must be continuously resident in Denmark for at least 10 years prior to their index-date to be included as a case in the study. Demographic and residency data for cases and controls were retrieved from the CPR.

At the index date, each case is matched to six controls from the general Danish population by sex, date of birth and municipality of residence. Each control must also have been continuously resident in Denmark for at least 10 years prior to the index-date. Additionally, the controls must be resident in Denmark until the second in-or outpatient contact of their case, to avoid immortality bias. Finally, the control must be free of IBD diagnoses prior to the index-date. The study population was followed until a censoring event including emigration, death, 31 December 2021 (end of study), or any diagnosis of IBD (for matched controls).

### 2.2 Statistical analysis

We analyse the association between IBD and the 1583 diseases at the three-character level in Chapters (I-XIX) of ICD-10 (e.g., M72 *Fibroblastic disorders*). The analysis is split into two diagnostic windows (pre-diagnosis and post-diagnosis) for each three-character disease category. First, we calculate the cumulative incidence spanning from 10 years prior up to 10 years after the IBD diagnosis date for the first appearance of any diagnosis by disease category within each chapter. We then calculate pre– and post-diagnosis risk of ICD-10 disease category diagnosis and finally, calculate risk of pre-diagnosis association compared with post-diagnosis association of statistically significant disease categories with large effect size.

The IBD pre-diagnostic period is defined as the nine-year period from ten years prior to one year before the index-date. We exclude the most recent year prior to diagnosis from the pre-diagnostic period to avoid capturing sequelae of IBD management following diagnosis in the pre-diagnostic period. For each disease category, we record the number of cases and controls with that diagnosis in the pre-diagnostic period, perform Fisher’s exact test to compute the p-value and calculate an odds ratio estimate (OR) with a confidence interval.

The post-diagnostic period is defined as the period immediately after diagnosis and ends at the point of censoring or ICD-10 disease category diagnosis. For each disease category, the study population comprises all IBD cases and controls without a prior diagnosis of the disease category in question. Therefore, all diagnoses within a disease category are the first for the case or the match. We perform Cox proportional regression analysis for ICD-10 disease category diagnosis to compute a hazard ratio (HR), 95% confidence interval and p-value.

To correct for multiple testing, we adjust our significance threshold (from p*<*0.05) with the Bonferroni correction ^19^. We test 6332 disease associations (1583 disease categories for both CD and UC, in the pre– and post-diagnostic window) resulting in a Bonferroni adjusted p-value of 7.90 *×* 10*^−^*^6^, which we refer to as the threshold for disease-wide statistical significance.

In the interest of identifying potential novel disease associations, comorbidities with disease-wide statistical significance (in either the pre-or post-diagnostic period) are further screened for prevalence threshold of at least 0.2% for assessment. We exclude categories with vague or catch-all terms such as “M89 *Other disorders of bone*”. Certain estimates could not be reported due to anonymity requirements and low case numbers, in these cases the intervals are left blank in the figures.

Finally, we identify comorbidities with strong pre-diagnostic effects, relative to their post-diagnostic effects. This analysis includes only highly significant pre-diagnostic (p*<* 0.001) disease categories, where the ratio of pre-to post-diagnostic effect is greater than 1.2-fold.

To understand the generalizability of our findings based on Denmark’s registries, we compare effect sizes for IBD associated comorbidities with those available through the Finnish electronic registry system (FinRegistry). ^20^ The effect estimates that we could obtain through this resource did not distinguish between the pre– and post-diagnostic windows, which is an important limitation, but it does allow us to validate estimates. For the purposes of computing comparable estimates, we did not distinguish between pre– and post-diagnostic windows in the validation subanalysis.

To identify heterogeneity in relationships between IBD and comorbidities in FinRegistry relative to the Danish registries, we use Breslow-Day’s test with Tarone’s correction ^21^ as implemented by the R package metafor. ^22^ Some FinRegistry estimates did not include counts for privacy reasons, so for these diseases we cannot assess heterogeneity. To identify systematic biases of the registries as to the size of odds ratios, we conduct a paired Wilcoxon signed rank test. ^23^

## 3 RESULTS

We identified 11,298 CD patients and 23,067 UC patients diagnosed with IBD in Denmark in the period from 1 January 2005 to 31 December 2021. Each patient was matched to six individuals from the general Danish population by sex, date of birth and municipality (n = 206,190) using the date of diagnosis as index date (Figure S1). Among IBD patients, 56.4% of CD patients and 51.7% of UC patients were women, 50.9% of CD patients and 40.3% of UC patients were diagnosed before age 40 (Table 1).

**Table 1:** Demographic baseline characteristics (age (years), year of diagnosis, sex at birth) of inflammatory bowel disease patients (Crohn’s disease and ulcerative colitis) and sex– and age-match controls.

|  | Crohn's disease |  | Ulcerative colitis |  |
| --- | --- | --- | --- | --- |
|  | N = 11,298 | controls, N = 67,788 | N = 23,067 | controls, N = 138,402 |
| **Age of IBD onset (index date), y** |  |  |  |  |
| 17-39 | 5,749 (50.9%) | 34,458 (50.8%) | 9,306 (40.3%) | 55,817 (40.3%) |
| 40-59 | 3,144 (27.8%) | 18,934 (27.9%) | 7,401 (32.1%) | 44,375 (32.1%) |
| 60+ | 2,405 (21.3%) | 14,396 (21.2%) | 6,360 (27.6%) | 38,210 (27.6%) |
| **Calendar year of birth** |  |  |  |  |
| 1905-1939 | 740 (6.5%) | 4,440 (6.5%) | 2,501 (10.8%) | 15,006 (10.8%) |
| 1940-1959 | 2,508 (22.2%) | 15,048 (22.2%) | 6,230 (27.0%) | 37,380 (27.0%) |
| 1960-1979 | 3,506 (31.0%) | 21,036 (31.0%) | 7,740 (33.6%) | 46,440 (33.6%) |
| 1980-2003 | 4,544 (40.2%) | 27,264 (40.2%) | 6,596 (28.6%) | 39,576 (28.6%) |
| **Calendar year of IBD diagnosis (index date)** |  |  |  |  |
| 2005-2009 | 2,724 (24.1%) | 16,344 (24.1%) | 6,977 (30.2%) | 41,862 (30.2%) |
| 2010-2014 | 3,261 (28.9%) | 19,566 (28.9%) | 7,265 (31.5%) | 43,590 (31.5%) |
| 2015-2018 | 3,134 (27.7%) | 18,804 (27.7%) | 5,494 (23.8%) | 32,964 (23.8%) |
| 2019-2021 | 2,179 (19.3%) | 13,074 (19.3%) | 3,331 (14.4%) | 19,986 (14.4%) |
| **Sex** |  |  |  |  |
| Female | 6,368 (56.4%) | 38,208 (56.4%) | 11,916 (51.7%) | 71,496 (51.7%) |
| Male | 4,930 (43.6%) | 29,580 (43.6%) | 11,151 (48.3%) | 66,906 (48.3%) |

### 3.1 Cumulative incidence of comorbidities pre– and post-IBD

We observed an overall pattern of multimorbidity across organ systems, beginning up to 10 years prior to diagnosis and being increasingly pronounced over time. We particularly observe a significantly increased risk of cardiometabolic, neuropsychological, and skin and musculoskeletal disorders present up to a decade before IBD diagnosis (Figure 1). For most organ systems, the excess occurrence of diseases both pre– and post-diagnostically was more pronounced in CD than in UC (Figure 1; Figure S2).

**Figure 1:**
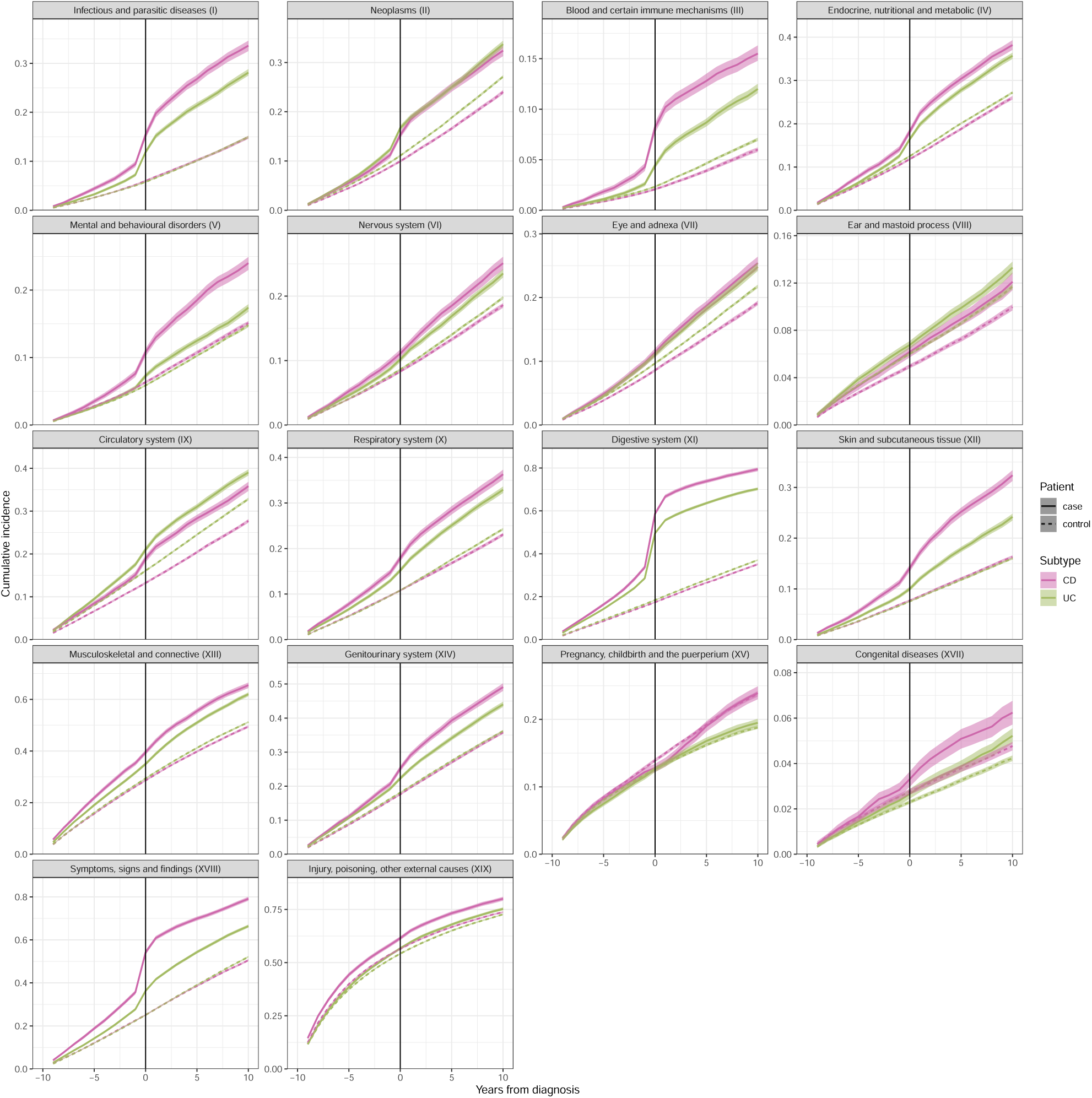
Cumulative incidence of three-character level diagnosis presented by ICD-10 chapter for CD, Crohn’s disease and UC, ulcerative colitis and age-, sex-, and municipality-matched controls from –10 years to 10 years from IBD, inflammatory bowel disease diagnosis.

Figures 2a and 2b illustrate the pre– and post-diagnostic risk of comorbidities for CD patients for ICD-10 chapter I to XI and XII to XIX, respectively. Whereas Figures 3a and 3b illustrate the pre– and post-diagnostic risk of comorbidities for UC patients for ICD-10 chapter I to XI and XII to XIX, respectively.

**Figure 2a:**
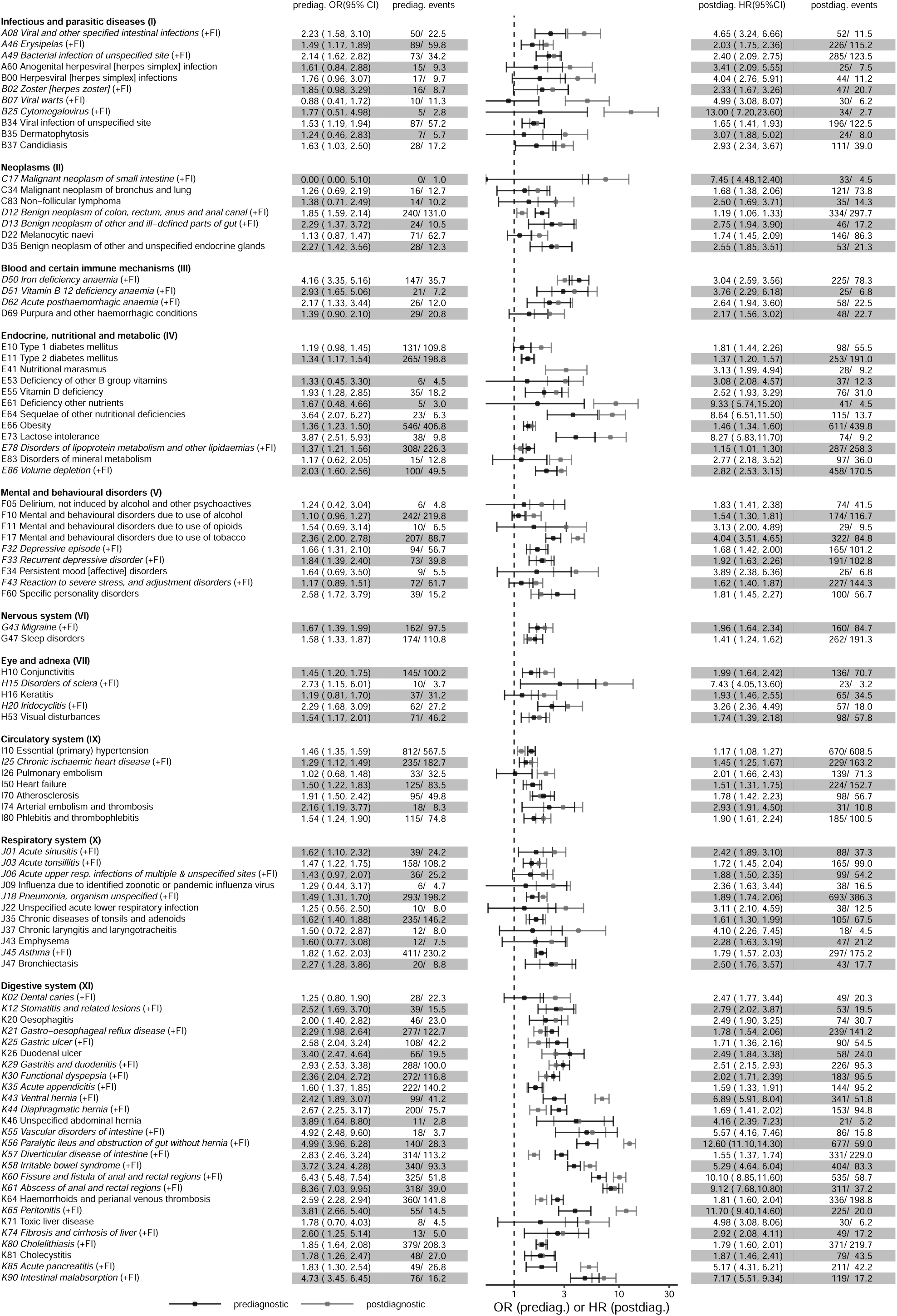
Crohn’s disease associated diagnoses at the ICD-10 three-character level, first part (Chapters I-XI). Pre-diagnostic OR, odds ratio and 95% CI, confidence intervals and post-diagnostic HR, hazard ratio and 95% CI estimates compared with age-, sex– and municipality-matched controls. Diagnoses meeting the prevalence threshold (*≥* 0.2%) with disease-wide significance (Bonferroni correction: p*<* 7.90 *×* 10*^−^*^6^) are included. The “(+FI)” symbol indicates that this effect is also significant in the Finnish registry.

**Figure 2b:**
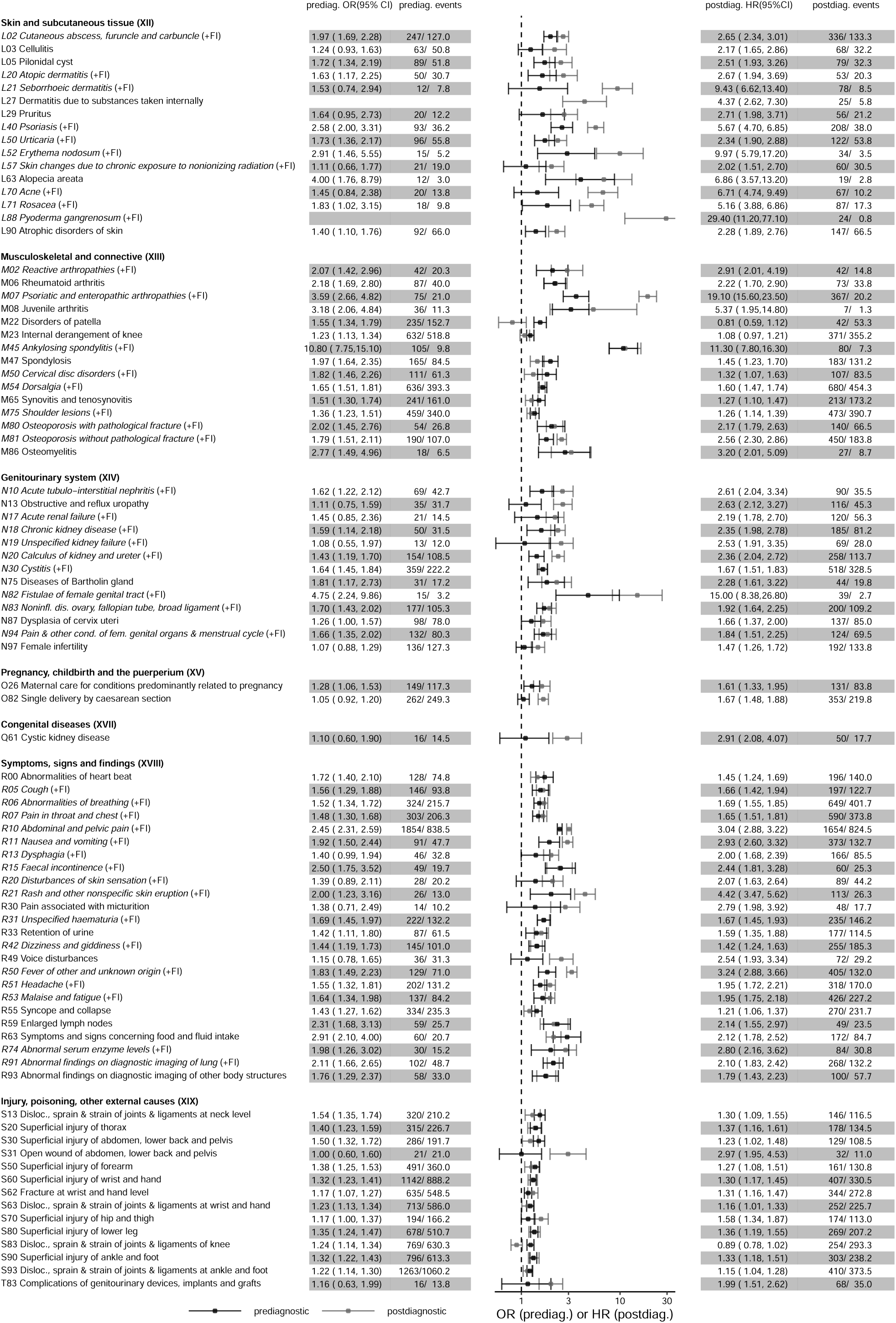
Crohn’s disease associated diagnoses at the ICD-10 three-character level, first part (Chapters XI-XIX). Pre-diagnostic OR, odds ratio and 95% CI, confidence intervals and post-diagnostic HR, hazard ratio and 95% CI estimates compared with age-, sex– and municipality-matched controls. Diagnoses meeting the prevalence threshold (*≥* 0.2%) with disease-wide significance (Bonferroni correction: p*<* 7.90 *×* 10*^−^*^6^) are included. The “(+FI)” symbol indicates that this effect is also significant in the Finnish registry.

**Figure 3a:**
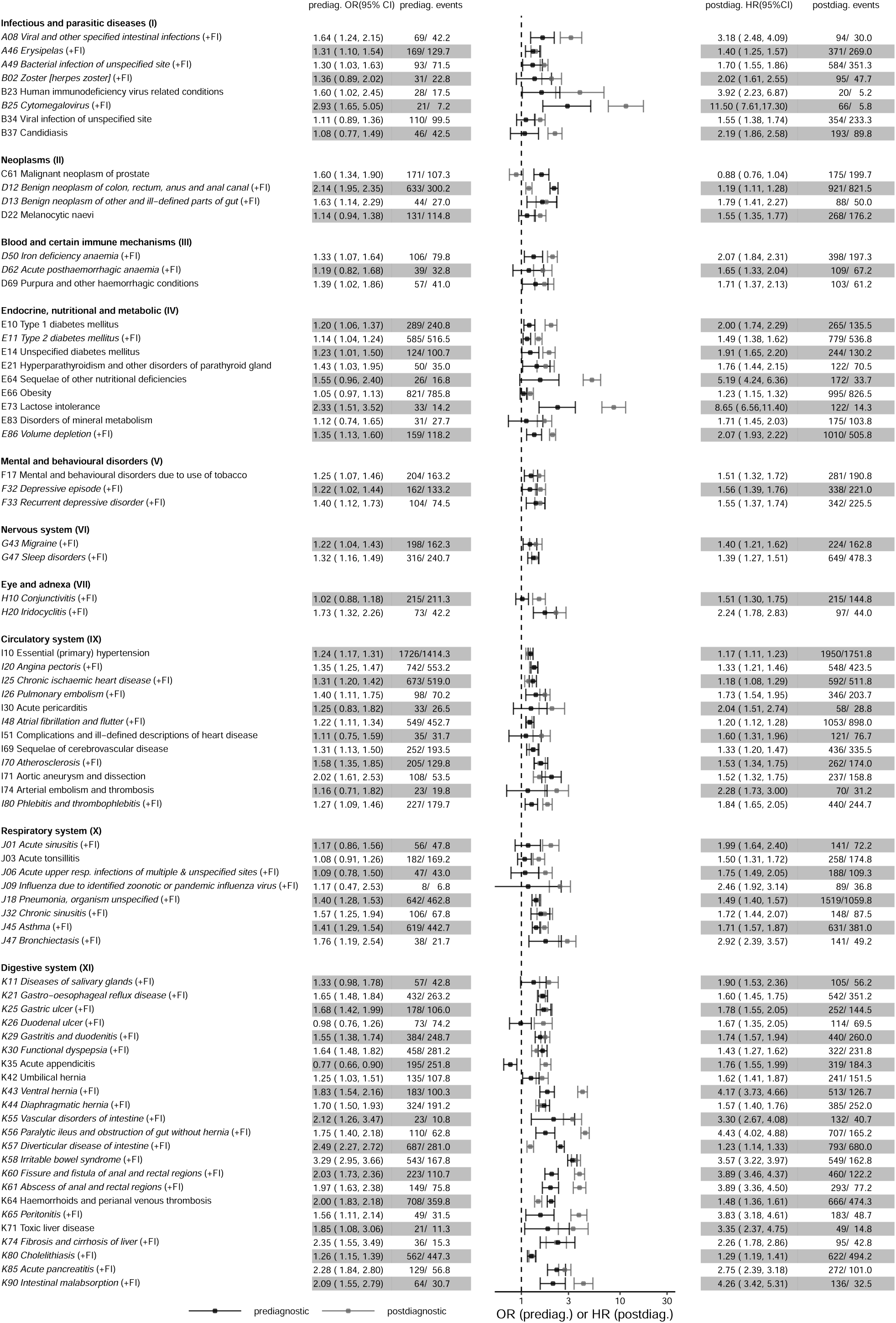
Ulcerative colitis associated diagnoses at the ICD-10 three-character level, first part (Chapters I-XI). Pre-diagnostic OR, odds ratio and 95% CI, confidence intervals and post-diagnostic HR, hazard ratio and 95% CI estimates compared with age-, sex– and municipality-matched controls. Diagnoses meeting the prevalence threshold (*≥* 0.2%) with disease-wide significance (Bonferroni correction: p*<* 7.90 *×* 10*^−^*^6^) are included. The “(+FI)” symbol indicates that this effect is also significant in the Finnish registry.

**Figure 3b:**
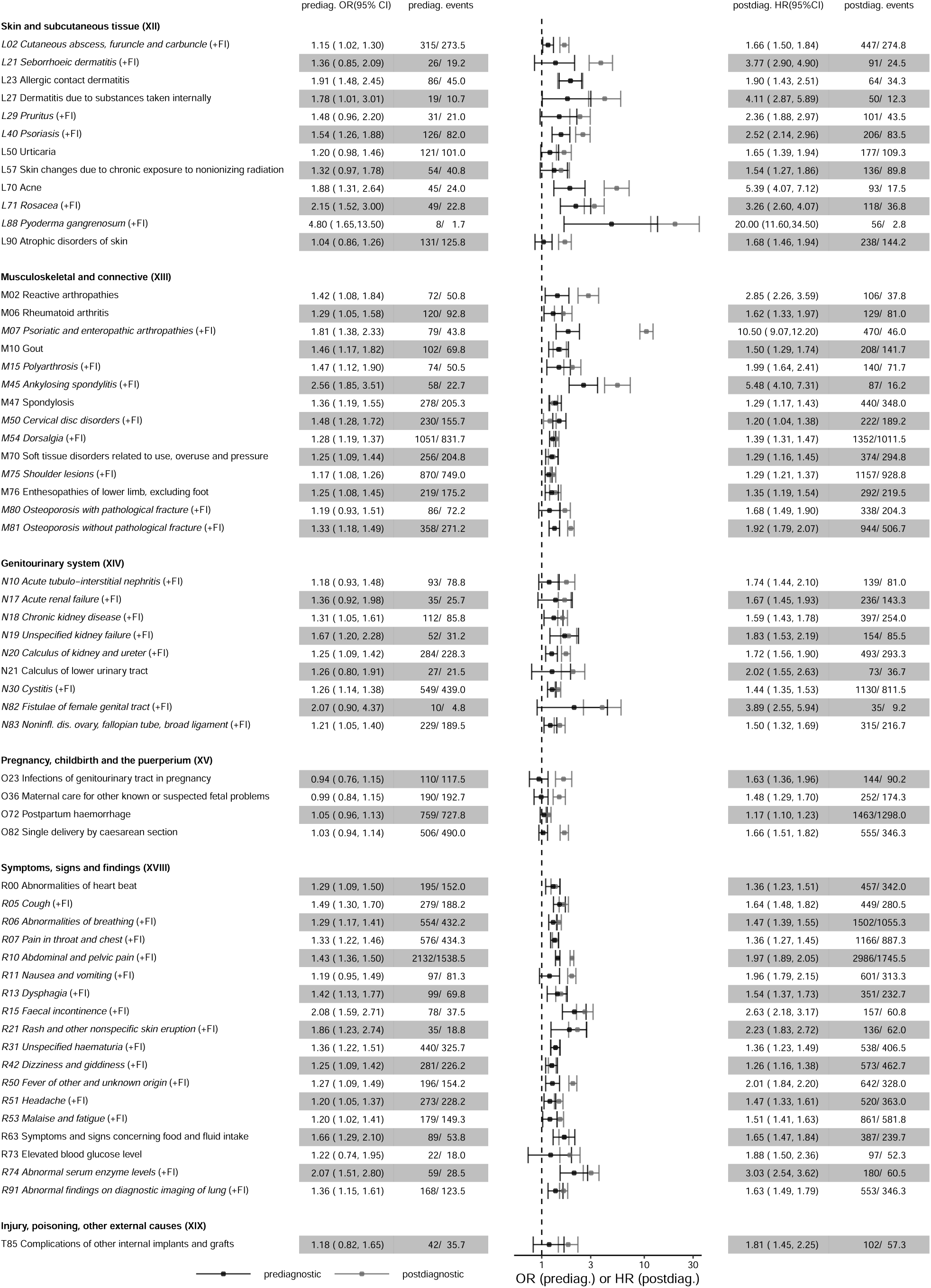
Ulcerative colitis associated diagnoses at the ICD-10 three-character level, first part (Chapters XI-XIX). Pre-diagnostic OR, odds ratio and 95% CI, confidence intervals and post-diagnostic HR, hazard ratio and 95% CI estimates compared with age-, sex– and municipality-matched controls. Diagnoses meeting the prevalence threshold (*≥* 0.2%) with disease-wide significance (Bonferroni correction: p*<* 7.90 *×* 10*^−^*^6^) are included. The “(+FI)” symbol indicates that this effect is also significant in the Finnish registry.

### 3.2 Pre– and post-diagnostic EIMs and IMIDs

We confirmed the occurrence of a series of EIMs after IBD diagnosis and were able to show that the majority of these EIMs already occurred in excess up to 10 years prior to IBD diagnosis, including diseases of the joints, eyes, skin, and respiratory tract. Joint and musculoskeletal disorders, such as reactive arthropathies, were seen at increased rates in both CD (OR: 2.07 95% CI: [1.42-2.96]) and UC (OR: 1.42 [1.08-1.84]) prior to IBD diagnosis, and even more pronounced after IBD diagnosis. Pyoderma gangrenosum was only increased prior to diagnosis in UC (OR_UC_: 4.80 [1.65-13.50]), despite a markedly excess risk in CD after diagnosis (HR_CD_: 29.40 [11.20-77.10]).

We confirmed an increased risk of multiple IMIDs in IBD patients versus the matched control population, and further showed that the increased risk was present up to 10 years prior to IBD diagnosis, as illustrated by estimates for psoriasis (OR_CD_: 2.58 [2.00-3.31]; OR_UC_: 1.54 [1.26-1.88]) and rheumatoid arthritis in CD (OR: 2.18 [1.69-2.80]).

### 3.3 Pre-diagnostic gastrointestinal and liver diseases

Several gastrointestinal and liver disorders, which were expected post-diagnostically, were already observed in excess in the pre-diagnostic period for both CD and UC. Diagnoses of chronic gastrointestinal diseases associated with IBD, including gastro-oesophageal reflux disease (OR_CD_: 2.29 [1.98-2.64]; OR_UC_: 1.65 [1.48-1.84]), gastric (OR_CD_: 2.58 [2.04-3.24]; OR_UC_: 1.68 [1.42-1.99]) and duodenal ulcers (OR_CD_: 3.40 [2.47-4.64]; OR_UC_: 0.98 [0.76-1.26]) showed stronger pre-diagnostic association with CD than UC. As expected, acute appendicitis prior to IBD diagnosis was associated with increased risk of CD (OR_CD_: 1.60 [1.37-1.85]) and decreased risk of UC (OR_UC_: 0.77 [0.66-0.90]).

### 3.4 Pre– and post-diagnostic cardiometabolic disorders

We observed an increased risk of cardiovascular diseases in the pre-diagnostic period, including hypertension (OR_CD_: 1.46 [1.35-1.59]; OR_UC_: 1.24 [1.17-1.31]) and ischemic heart disease (OR_CD_: 1.29 [1.12-1.49]; OR_UC_: 1.31 [1.20-1.42]) for both CD and UC, and arterial embolism and thrombosis (OR_CD_: 2.16 [1.19-3.77]) in CD, which increased over time in the post-diagnostic period. We furthermore observed an increased risk of type 2 diabetes mellitus both prior to (OR_CD_: 1.34 [1.17-1.54]; OR_UC_: 1.14 [1.04-1.24]) and after IBD diagnosis, as well as an increased risk of obesity, particularly in the post-diagnostic period (HR_CD_: 1.46 [1.34-1.6]; HR_UC_: 1.23 [1.15-1.32]).

### 3.5 Pre– and post-diagnostic neuropsychological disorders including substance use

Excess risk of several mental and behavioural disorders, including depression, anxiety, and substance use disorders was observed in the pre– and post-diagnostic period for CD patients in particular, who had a markedly increased risk of alcohol (HR: 1.54 [1.30-1.81]), tobacco use disorders (HR: 4.04 [3.51-4.65]), and stress and adjustment disorders (HR: 1.62 [1.40-1.87]) post-diagnostically.

All effect sizes calculated for comorbidities associated with CD and UC in the pre– and post-diagnostic period are presented in Table S1.

### 3.6 Disease risk in pre-diagnostic relative to post-diagnostic phase

Finally, we assessed which diseases were more prominent in the *pre-diagnostic* relative to the *post-diagnostic* period (Figure 4). Overall, there were more diseases occurring predominantly pre-diagnostically, compared to the post-diagnostically in patients with CD (14 diseases) than in patients with UC (10 diseases), confirming the more pronounced systemic prodromal development of CD and include expected diagnoses, such as haemorrhoids (OR: 2.59 [2.28-2.94]; HR: 1.81 [1.60-2.04]), as well as unexpected diagnoses, such as hypertension which was more prominent prior to diagnosis (OR: 1.46 [1.35-1.59]; HR: 1.17 [1.08-1.27]). Lastly, UC patients were at higher risk of being diagnosed with stress before compared with after UC diagnosis (OR: 1.41 [1.17-1.70]; HR: 1.08 [0.95-1.22]), potentially reflecting delayed diagnosis of IBD reflected by increased symptoms of deterioration.

### 3.7 Meta-analysis

To test our findings, we looked up all codes, presented in Figures 2-4 in the Finnish data (with the limitation that the Finnish estimates did not distinguish between the pre-diagnostic and post-diagnostic periods) and marked all estimates in Figures 2-4 with (+FI) if our associations were confirmed in the Finnish data. Further, direct comparison of Danish and Finnish effect sizes are shown in Figure 5, where we also indicate where there is evidence of inconsistency between Finland and Denmark, when corrected for multiple testing. Effects which were stronger in Denmark appear above the line, and effects which were stronger in Finland appear below the line.

**Figure 4:**
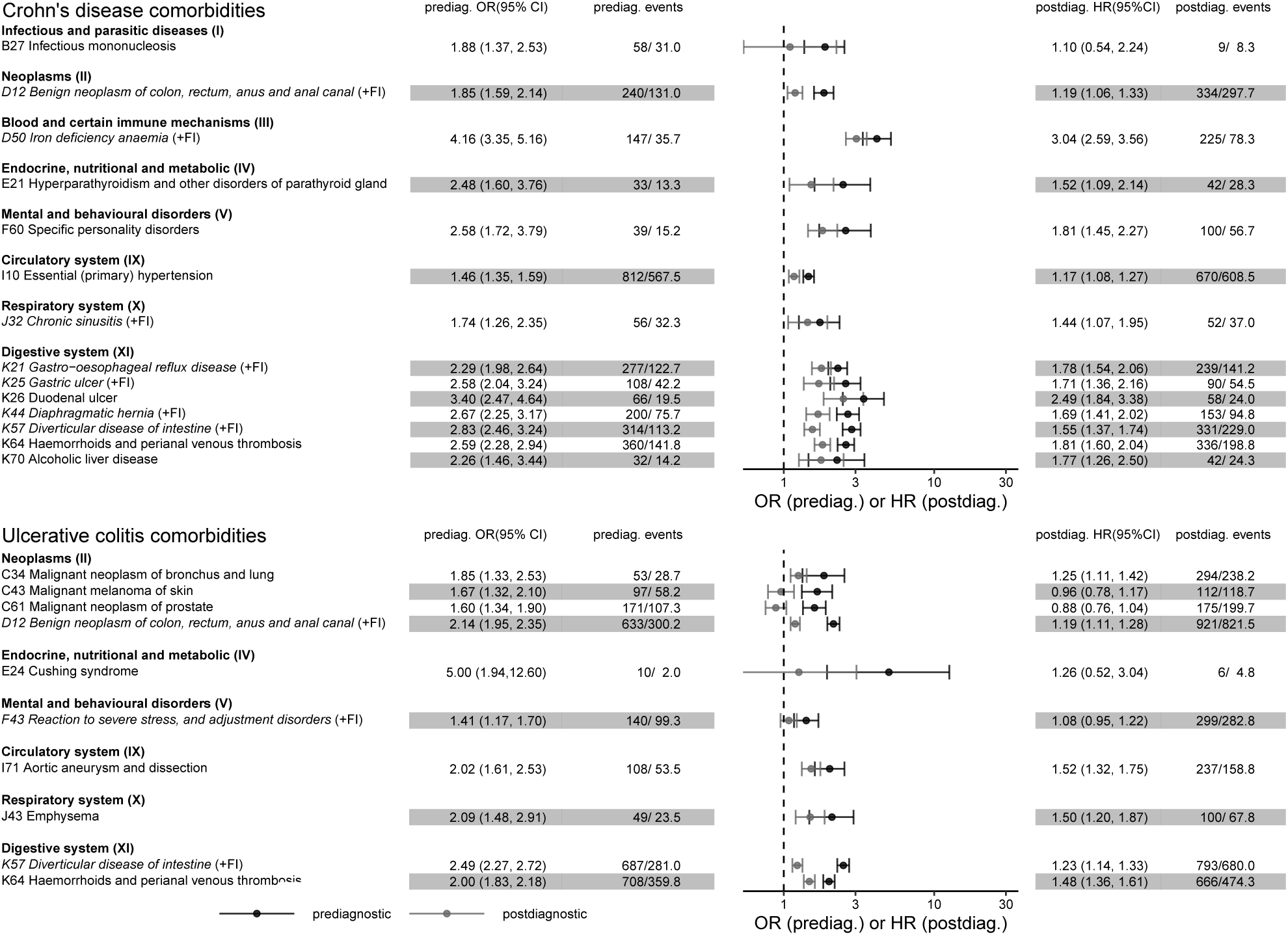
Diagnoses associated with CD, Crohn’s disease and UC, ulcerative colitis before IBD, inflammatory bowel disease diagnosis. Pre-diagnostic OR, odds ratio compared with post-diagnostic HR, hazard ratio of diagnoses of disease-wide significance for Crohn’s disease and Ulcerative colitis. Highly statistically significant diagnoses (p *<* 0.001), with pre-diagnostic effect estimates (OR) at least 1.2 of the post-diagnostic effect estimates (HR) are included. The “(+FI)” symbol indicates that this effect is also significant in the Finnish registry.

**Figure 5:**
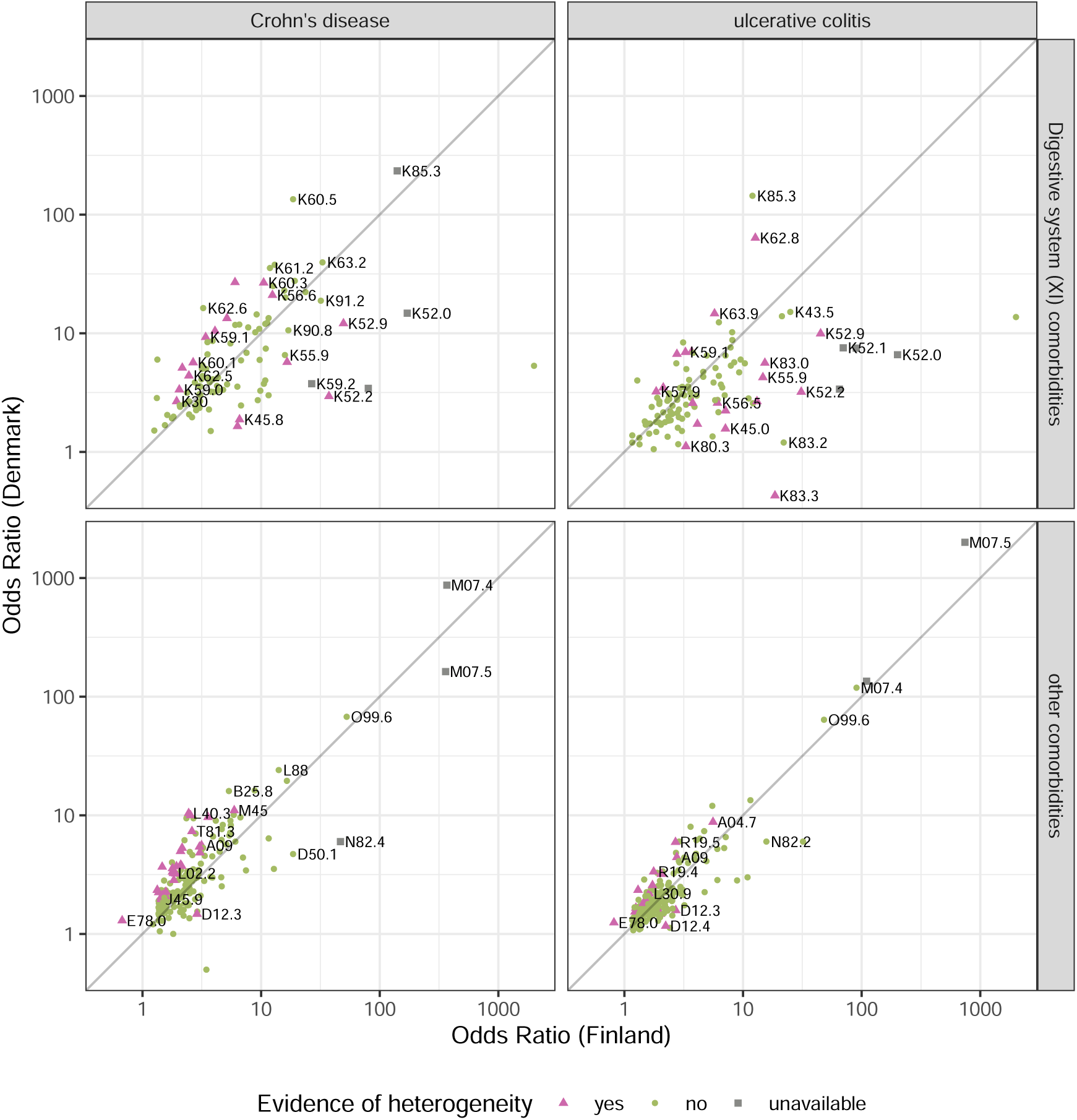
Comparison of effect sizes for non-digestive comorbidities in Crohn’s (left) and ulcerative colitis (right) as reported by Risteys (Finland) compared to the odds ratios computed from the Danish registry. Points beyond 1000 have estimates which are infinite in the relevant axis, but are included here for completeness.

There was evidence of heterogeneity of ORs between the two countries in only 15.7% (44/281) and 11.4% (35/301) of diseases in CD and UC respectively. In every disease except one, the directions of the effect size were consistent. The one exception was E78.0 Pure hypercholesterolaemia, where the Finnish data indicated a protective effect for IBD (OR_CD_: 0.67 [0.60-0.67]; OR_UC_: 0.82 [0.76-0.88]), whereas IBD was a risk factor in Denmark (OR_CD_: 1.30 [1.18-1.42]; OR_UC_: 1.25 [1.18-1.32]). We conducted a paired Wilcoxon signed rank test between the registry systems. Crohn’s disease ORs were systematically higher in Denmark relative to Finland (p*<* 1 *×* 10*^−^*^6^), whereas there was no significant difference between effect sizes for Danish and Finnish patients with UC (p = 0.203).

## 4 DISCUSSION

This comprehensive population-based disease-wide association study examining *>*1500 diseases among *>*35,000 patients with IBD and *>*200,000 controls revealed that a diagnosis of IBD was associated with 312 other diseases, of which 125 diseases were present up to 10 years before diagnosis. Based on these observations, we created an interactive resource to explore IBD comorbidities across organ systems (Table S1). In addition to identifying expected diseases associated with IBD, including known EIMs and IMIDs, we identified multisystem comorbidities up to a decade prior to the incident IBD diagnosis, including cardiometabolic, neuropsychological, and immune and infectious diseases associated with both CD and UC. Most of the pre-diagnostic comorbidities appeared with larger effect sizes and earlier in CD than UC patients. Results were validated and confirmed with respect to publicly available (albeit cruder) Finnish estimates.

The validity of our findings is further confirmed by observation of an expected increased risk of EIMs in patients with IBD, including erythema nodosum, asthma, pyoderma gangrenosum, and iridocyclitis, as well as an increased risk of IMIDs, such as psoriasis, ankylosing spondylitis and coeliac disease. ^24^ In addition to showing the exact magnitude of these risks at a population level representing the true spectrum of IBD (from the mildest to the most severe case), hence providing estimates that can be used for information of newly diagnosed patients with a yet unknown disease course, we also show that these disorders are present at significantly increased frequency several years before diagnosis, pointing towards a long prodromal phase of IBD with multiorgan involvement. We observe that the risk of EIMs and IMIDs in the post-diagnostic period is larger in CD than in UC, again highlighting CD particularly as a multisystem disease. Our findings may have implications for screening and therapeutic considerations and raises the question, whether EIM and IMID diagnoses should prompt practitioner investigation for IBD in the context of gastrointestinal symptoms.

In addition to EIMs and IMIDs, we observe an increased risk of cardiometabolic disorders following IBD diagnosis in accordance with former observational studies ^25;26^. We show that these diseases may be present up to a decade prior to IBD diagnosis, which may have both aetiological and treatment implications. The pre-IBD presence of cardiometabolic diseases indicates that not only chronic systemic inflammation, but also shared underlying mechanisms for development of cardiometabolic disease and IBD may exist, potentially reflecting shared environmental factors such as smoking and diet, which may act through microbiome dysfunction ^3^. The increased risk of cardiometabolic disease in the pre-diagnostic period may also have clinical implications. The risk of ischaemic heart disease is increased markedly prior to IBD diagnosis and warrants clinical awareness following an IBD diagnosis. As JAK inhibitors – a class of small oral molecules to treat chronic inflammatory diseases – have recently been linked with increased MACE (major adverse cardiovascular events) risk–in rheumatoid arthritis populations ^27^, additional caution might also be warranted in selected IBD patients in light of our observed pre-diagnostic cardiovascular comorbidity.

Regarding the psychiatric disorders, we confirm significant bidirectional association between IBD in relation to depression and anxiety ^28^. The proposed mechanisms underlying this association ^7^ are complex but may involve direct biochemical pathways through “the gut–brain axis” via the vagus nerve ^29^, or more indirect mechanisms, such as the, possibly preclinical, symptoms of IBD inducing psychological stress ^30^.

For several diseases primarily observed in excess following an IBD diagnosis, it remains to be understood if these are consequences of severe disease or the treatment thereof. Two recent meta-analyses ^31;32^ have demonstrated the harms associated with placebo — and hence untreated active IBD — in randomized controlled trials, but consequences of treatment still merit attention. We observed a significantly increased risk in the post-diagnostic period of osteopenia and osteoporosis with pathological fracture, which is a well-documented adverse effect of long-term steroid exposure. ^33^ Advances in IBD therapies over the last decades with more steroid sparing management in severe IBD management may change this picture over time. In addition to active disease, immunosuppression may explain the viral enteric, pulmonary, and opportunistic infections seen at higher rates in the post-compared with pre-diagnostic period. Balancing the adverse impact of therapies is an ongoing clinical challenge, and there is also a need for development of tools to differentiate between severity of disease and consequences of treatment thereof when assessing long-term outcomes with observational data.

The Danish LPR and FinRegister are shown to be broadly consistent regarding the computed effect sizes of IBD on other comobidities. Heterogeneity in effect sizes is to be expected since there are country-level differences in the age-structure and censoring distributions.

The strengths of the present study are the population-wide and disease-wide assessment of all potential known and unknown comorbidities of IBD, which to our knowledge is the first phenomic study of this nature for any chronic illness. Further strengths are the size of the case population, assuring power; the drawing of matched controls from the general population rather than non-IBD hospital records, improving the generalizability of our findings; the length of follow-up beginning from 10 years prior to diagnosis until up to 17 years post-diagnosis, allowing us to investigate long-term associations in either direction of diagnosis; and the termination of the pre-diagnostic window 1 year prior to the incident diagnosis of IBD, thereby reducing surveillance bias. Lastly, both our application of a prevalence threshold of 0.2% for disease assessment and our choice of the Bonferroni correction, leading to a disease-wide significance threshold of 7.90 *×* 10*^−^*^6^, reduces the potential for false positive findings.

Potential limitations that should be acknowledged include that registry data is subject to surveillance bias or capture of systemic biases such as common incorrect diagnoses by clinical practitioners, and this may have impacted our identification of several disorders diagnosed via e.g., endoscopy, including benign intestinal tumours or diverticular disease. Although we capture most EIMs and IMIDs associated with IBD and, as a novel finding, show that they appear many years prior to disease diagnosis, not all known IMIDs related to IBD will be captured, if not captured through outpatient or inpatient contacts. The study also contains risk of ascertainment bias, but this is be diminished by use of population-representative, nationwide diagnostic data, where cases are additionally matched to controls within municipality of residence. This is particularly the case for our findings in analyses from the pre-diagnostic period, where IBD patients would not be subject to the same level of clinical follow-up as following diagnosis.

In conclusion, in this population-based disease-wide study of almost 35,000 IBD patients followed for *>*300,000 pre-diagnostic and *>*250,000 post-diagnostic person-years (average 7.68 person-years per case), we observe an overall pattern of multimorbidity beginning from 10 years prior to diagnosis, with a more pronounced effect in CD relative to UC. Findings indicate the presence of multisystem pathology, which is increasing in frequency throughout the disease course. This underscores the need for multidisciplinary care, including cardiometabolic measurements and neuropsychological support, and increased attention to the management of a broad range of co-occurring disorders already present at the time of IBD diagnosis.

## Supporting information

Table S1: Interactive table of effect sizes.

## Data Availability

The study was based on data from the Danish National Health registers (https://sundhedsdatastyrelsen.dk). The register data are protected by the Danish Act on Processing of Personal Data and are accessed through application to and approval from the Danish Data Protection Agency and the Danish Health Data Authority.

## Author contributions

ACE, GP, MB, and TJ conceptualised the paper and RE contributed key analysis concepts. All authors were involved in the developing the methodology, and validation of findings. ACE and GP developed the code for and undertook the analysis. ACE, GP, and MB provided the formal statistical methodology input. AS and ACE developed the Finnish validation analysis. The original draft was written by RE, ACE, MB, CWL, and TJ. All authors contributed to review and editing of the manuscript. ACE, RE, MB, BV, CWL, AS and TJ contributed to the visualisation of findings.

## Acknowledgements

We wish to thank Carsten Eriksen for his assistance with improving the readability of the forest plots.

## Supplementary materials

**Table S1:**
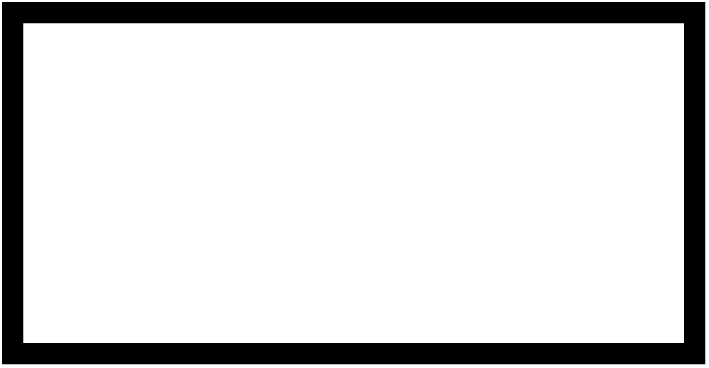
Interactive table of effect sizes. **See online supplementary materials.**

**Figure S1:**
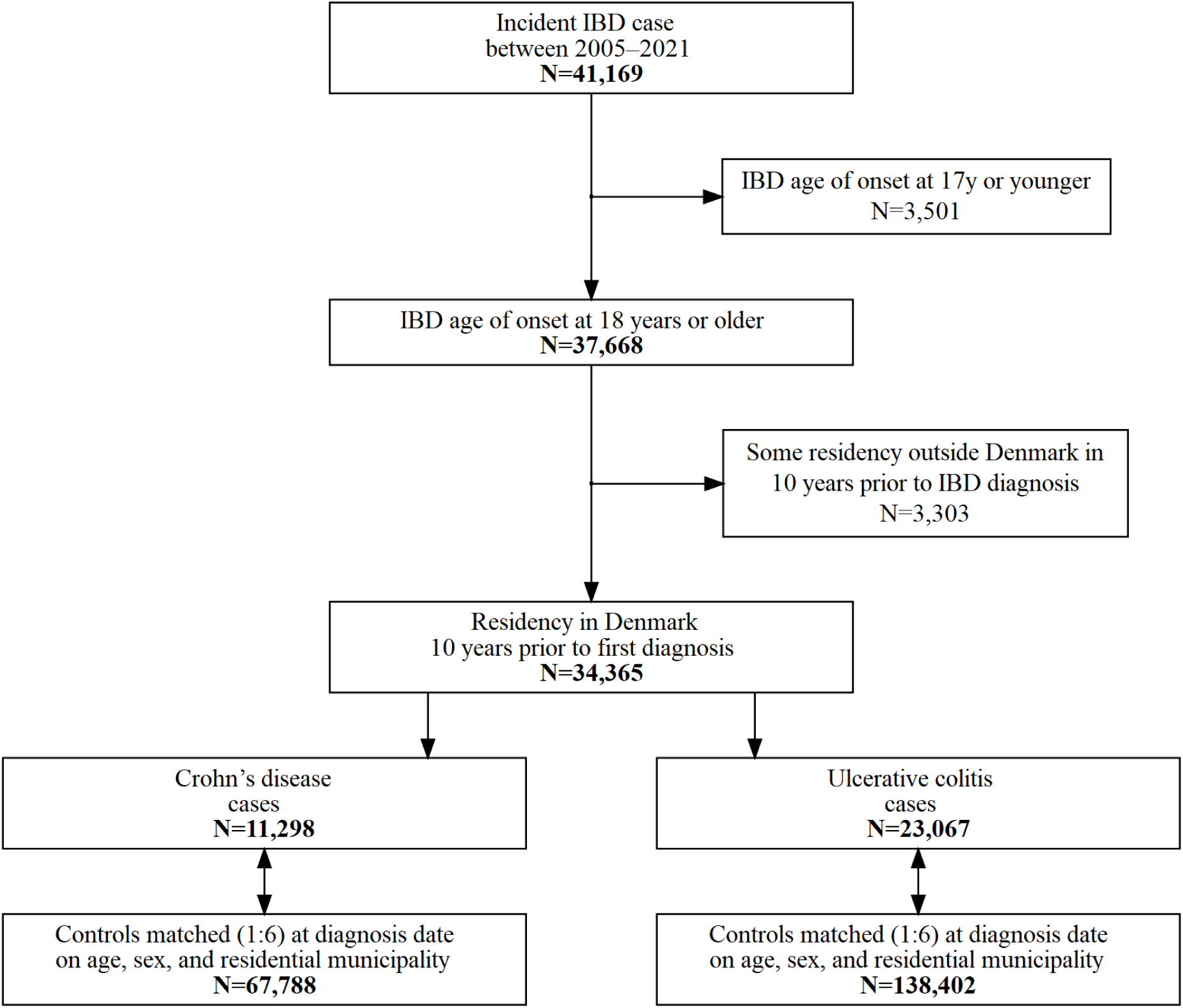
Flow diagram of study population.

**Figure S2:**
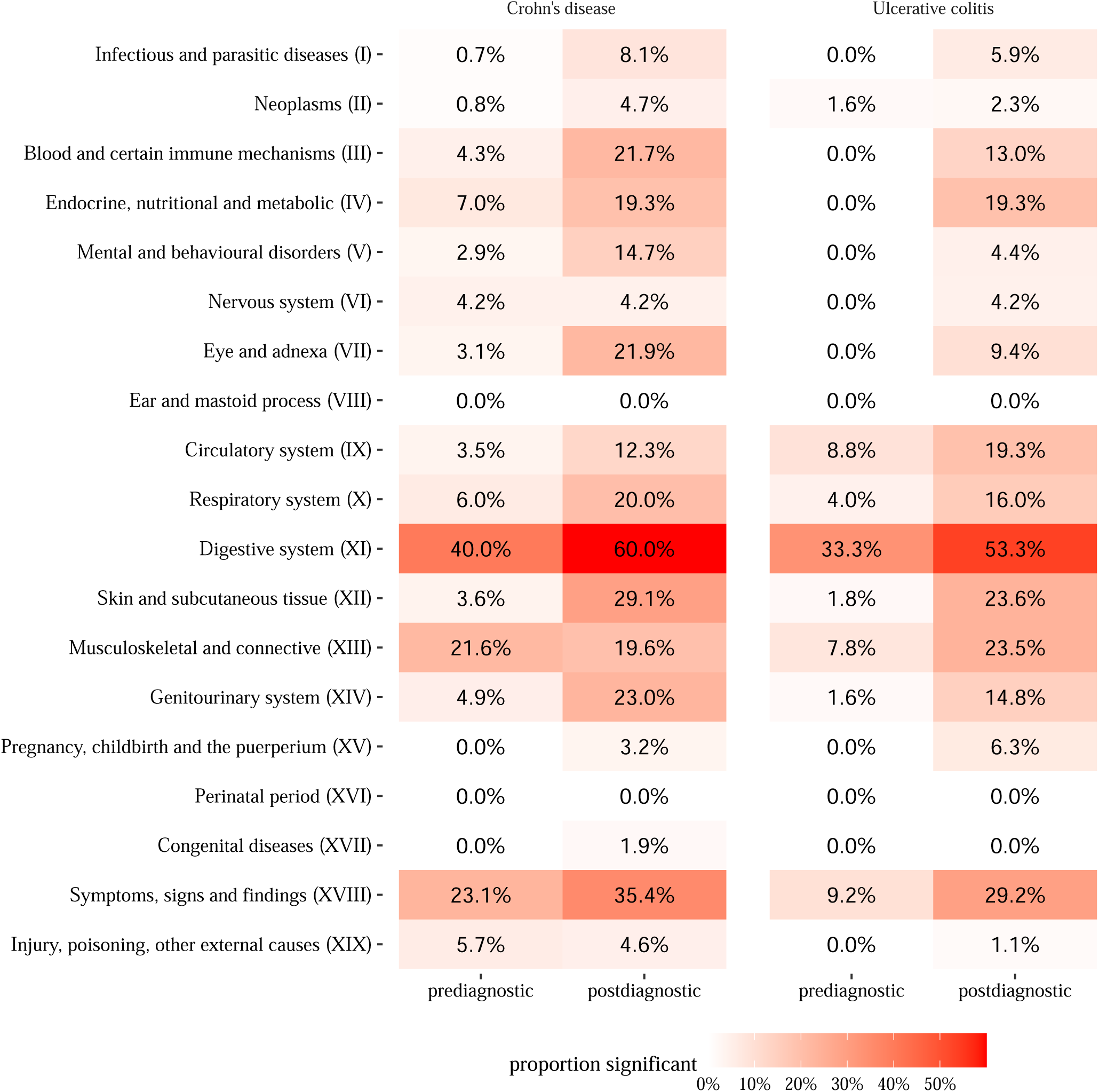
Proportion of significant associations in each ICD-10 Chapter. Significance threshold corrected for multiple testing.

